# IMPACT - Phase Ib Trial of Intramuscular Personalized Neoantigen Synthetic Long Peptide Vaccines in Patients with Advanced Melanoma and Renal Cell Carcinoma

**DOI:** 10.64898/2025.12.02.25341434

**Authors:** Nussara Pakvisal, Piriya Wongkongkathep, Worawan Bunrasmee, Pimpayao Sodsai, Jirattha Siriluksana, Nittaya Boonnak, Tawanchay Sangcharoen, Bussaba Trakarnsanga, Shama Sukprakun, Peepattra Wantanasiri, Korn Chotirosniramit, Meghna Phanichkrivalkosil, Saharat Nanthawong, Prangwalai Chanchaem, Suwanan Mankhong, Sarinya Kumpunya, Suangson Supabphol, Nitiwat Sirijun, Krittanon Kongtragulsub, Phorutai Pearngam, Poorichaya Somparn, David Michael Payne, Albert Reynolds, Boyang Zhao, Verayuth Praphanphoj, Natapol Pornputtapong, Sira Sriswasdi, Duangdao Wichadakul, Suleepon Uttamapinan, Pattama Angspatt, Ploytuangporn Wongchanapat, Nattaya Teeyapun, Sutima Luangdilok, Piyada Sitthideatphaiboon, Thiti Susiriwatananont, Nicha Zungsontiporn, Napa Parinyanitikul, Suebpong Tanasanvimon, Chanida Vinayanuwattikun, Andres Salazar, Nattiya Hirankarn, Virote Sriuranpong, Trairak Pisitkun

**Affiliations:** Division of Medical Oncology, Department of Medicine, Faculty of Medicine, Chulalongkorn University and The King Chulalongkorn Memorial Hospital, Bangkok, Thailand; Center of Excellence in Systems Biology, Faculty of Medicine, Chulalongkorn University, Thailand; Center of Excellence in Immunology and Immune-Mediated Diseases, Faculty of Medicine, Chulalongkorn University, Thailand; Maha Chakri Sirindhorn Clinical Research Center, Faculty of Medicine, Chulalongkorn University, Thailand; Pharmacy Department, King Chulalongkorn Memorial Hospital, Thailand; Department of Biochemistry, Faculty of Medicine, Chulalongkorn University, Thailand; Center of Excellence in Systems Microbiology, Faculty of Medicine, Chulalongkorn University, Thailand; Department of Physiology, Faculty of Medicine, Khon Kaen University; Faculty of Medicine, Prince of Songkla University; School of Medicine, Vanderbilt University, USA; Quantalarity Research Group, USA; Medical Genetics Center, Thailand; Department of Biochemistry and Microbiology, Faculty of Pharmaceutical Sciences, Chulalongkorn University, Thailand; Center of Excellence in Computational Molecular Biology, Faculty of Medicine, Chulalongkorn University, Thailand; Department of Computer Engineering, Faculty of Engineering, Chulalongkorn University, Thailand; Medical Oncology Unit, The King Chulalongkorn Memorial Hospital, Thailand; Wattanosoth Cancer Hospital, Thailand; Oncovir, Inc., USA

## Abstract

**Purpose:** To evaluate safety and immunogenicity of intramuscularly delivered personalized neoantigen synthetic long peptide (SLP) vaccines in patients with advanced solid tumors.

**Patients and Methods:** In this Phase I trial, 12 patients with advanced melanoma (n=9) or renal cell carcinoma (n=3) who could no longer access further standard treatments received intramuscular neoantigen SLP vaccines with poly-ICLC. Each vaccine contained ∼20 predicted neoantigen peptides. Adverse events were monitored throughout vaccination and follow-up. Immune profiling was performed at baseline and predefined post-vaccination time points.

**Results:** Intramuscular neoantigen vaccination was well tolerated, with only grade 1–2 local pain or fever and no immune-mediated toxicities. All participants developed de novo T-cell responses, detectable within one week. On average, 46% of peptides per patient were immunogenic, inducing both CD8⁺ and CD4⁺ neoantigen-specific responses. Patients previously treated with immune checkpoint inhibitors (ICIs) had higher baseline immunity but achieved comparable post-vaccination responses to ICI-naïve patients. IFN-γ–dominant CD8⁺ and TNF-α–dominant CD4⁺ responses were observed, along with increased effector memory differentiation. Two patients with higher CD8⁺ TEMRA proportions were the longest survivors. Tumor biopsies revealed enhanced CD8⁺ infiltration, and epitope spreading occurred in one of two evaluable cases. Analysis of 239 peptides showed greater immunogenicity for dual MHC I/II–binding, cysteine-containing, and in-frame indel- or low-VAF–derived mutations, while proline substitutions reduced responses.

**Conclusions:** Intramuscular neoantigen SLP vaccination with poly-ICLC is safe and induces rapid, mutation-specific T-cell immunity with robust CD8⁺ effector responses. These findings support intramuscular administration as a promising strategy for peptide-based cancer vaccines.

**Translational relevance:** Personalized neoantigen vaccines offer a promising strategy to enhance tumor-specific immunity, but most prior studies using intradermal or subcutaneous delivery have shown limited induction of cytotoxic CD8⁺ T cells. This study demonstrates that intramuscular administration of personalized neoantigen synthetic long peptide vaccines with poly-ICLC is safe, feasible, and capable of eliciting rapid, mutation-specific CD4⁺ and CD8⁺ T-cell responses in patients with advanced melanoma and renal cell carcinoma. Vaccine-induced immunity was dominated by IFN-γ–producing cells and accompanied by a shift toward effector memory phenotypes. In selected cases, post-treatment tumor biopsies revealed increased CD8⁺ infiltration. These findings support intramuscular delivery as a practical and effective platform for neoantigen-based cancer vaccine.

## Introduction

Immune checkpoint inhibitors (ICIs) have transformed treatment landscape for multiple cancers, yet a substantial proportion of patients either fail to respond or ultimately develop resistance [1, 2]. These limitations has prompted the development of complementary immunotherapeutic strategies capable of generating robust, tumor-specific T cell responses. Personalized cancer vaccines targeting neoantigens, which are tumor-specific antigens derived from somatic mutations, offer a promising approach to induce focused antitumor immunity while minimizing off-target toxicity and risk of autoimmunity [3, 4]. Several vaccine platforms are under investigation, including mRNA [5, 6], DNA vectors [7], dendritic cell formulations [8–10], and synthetic long peptides (SLPs) [11, 12]. Among these, SLPs are particularly attractive due to their safety, biochemical stability, cost-effectiveness, and ability to deliver precisely epitopes with flexible adjuvant uses [13–15].

Recent clinical trials have demonstrated the feasibility and safety of personalized neoantigen peptide vaccines combined with poly-ICLC, a TLR-3 agonist, across various malignancies, including melanoma and renal cell carcinoma (RCC), and other solid tumors [11, 12, 16, 17]. These studies consistently reported the induction or expansion of neoantigen-specific T cells. However, most studies have used subcutaneous or intradermal administration, which predominantly elicites CD4+ T cell responses. To enhance CD8+ T cell responses, Montanide has been incorporated to improve antigen availability [18]. Preclinical studies of poly-ICLC indicate that the route of delivery critically influences immunogenicity [19]. While intravenous (IV) and intramuscular (IM) administration of poly-ICLC appear to be more effective approaches for inducing cytotoxic CD8+ T cell responses than subcutaneous routes, IV delivery is limited clinically due to systematic toxicity concerns. In contrast, IM delivery, a widely established and well-tolerated route in vaccinology, represents another practical delivery approach. In murine models, IM injections elicited stronger cytotoxic CD8+ T cell responses compared with subcutaneous administration [19], likely due to the higher vascularity of muscle tissue facilitating antigen dispersion into circulation. Collectively, these findings provide a strong rationale for evaluating IM administration in cancer neoantigen peptide vaccines to activate both CD4+ and CD8+ T cells.

To directly assess the intrinsic safety and immunogenic activity of intramuscular SLP neoantigen vaccination, we conducted a Phase Ib clinical trial (IMPACT – IntraMuscular Personalized neoAntigen Cancer Therapy). This study investigated SLP-based neoantigen vaccines as monotherapy administered intramuscularly in both ICI-pretreated and ICI-naïve patients with advanced melanoma or RCC who no longer had access to standard therapies.

## Materials and Methods

### Study Design and Participants

This Phase Ib, single-center clinical trial was conducted at the King Chulalongkorn Memorial Hospital, Bangkok, Thailand. Twelve patients with metastatic or unresectable melanoma or RCC who had progressed on or had no access to standard treatments were enrolled between January 2021 and June 2023 (Figure 1A). Eligible patients were required to have an Eastern Cooperative Oncology Group (ECOG) performance status of 0 or 1. Patients with prior treatment and progression on ICIs were allowed. Key exclusion criteria included active autoimmune disease and active infection.

**Figure 1:**
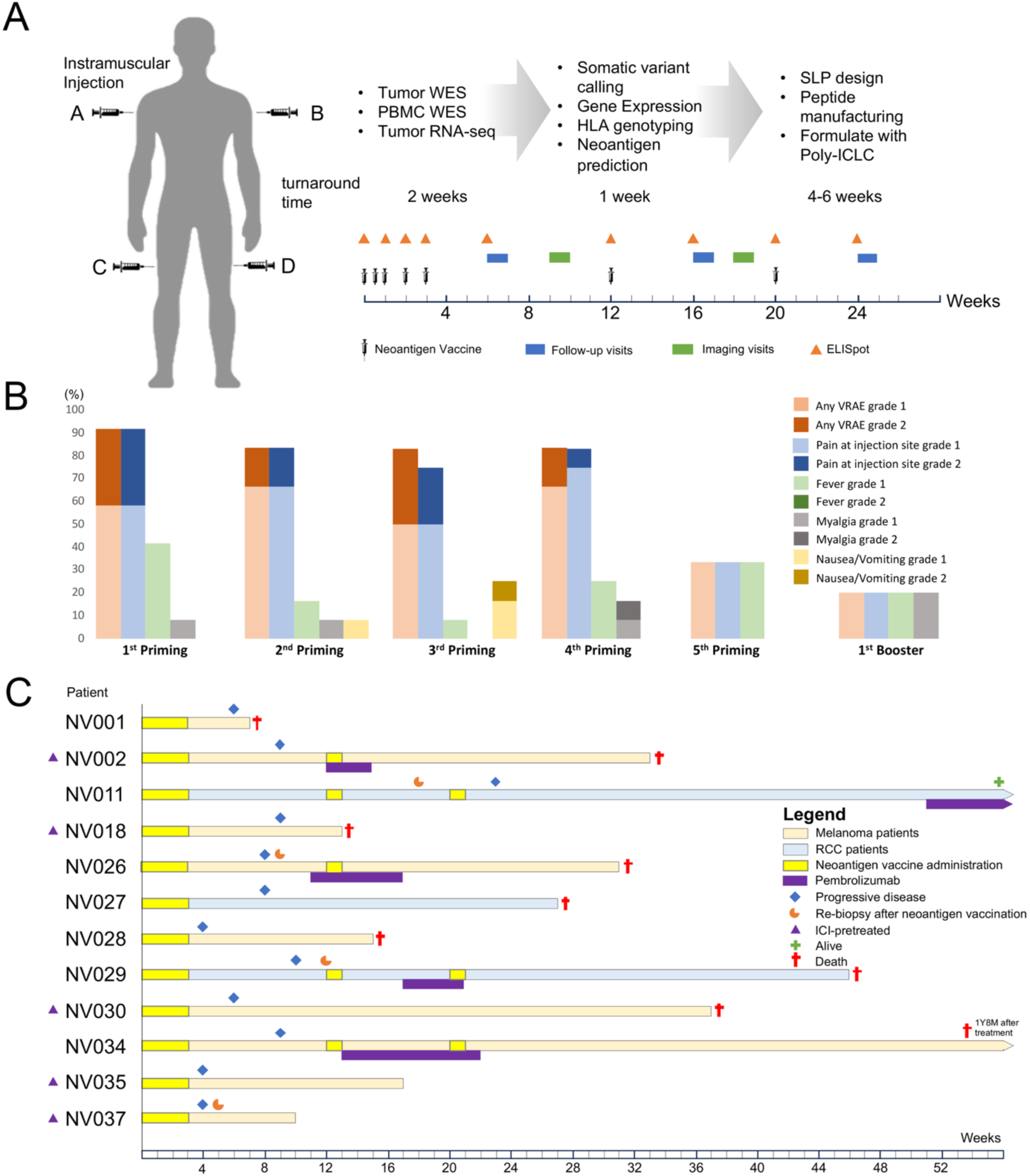
Trial design, study workflow, patient journey, clinical outcomes, safety and tolerability of the neoantigen vaccine. (A) Schematic overview of the personalized neoantigen vaccine trial. The process includes tumor and blood sample collection, whole-exome and RNA sequencing, bioinformatic analysis for neoantigen prediction, and manufacturing of synthetic long peptides (SLPs). The clinical timeline illustrates the vaccination schedule, consisting of five priming doses and two boosters, along with time points for immunological monitoring by ELISpot, clinical follow-up visits, and tumor imaging assessments. (B) Summary of treatment-related adverse events (AEs) observed during the five priming doses (P1-P5) of the vaccination schedule. All AEs were grade 1-2. The stacked bar charts show the number of patients experiencing each type of AE at each dose, with the most common being fever, myalgia, and injection site pain. Summary of treatment-related adverse events (AEs) observed during the five priming doses (P1-P5) of the vaccination schedule. All AEs were grade 1-2. The stacked bar charts show the number of patients experiencing each type of AE at each dose, with the most common being fever, myalgia, and injection site pain. (C) Swimmer plot summarizing the treatment course and clinical outcomes for each of the 12 patients. Each bar represents one patient, with colors indicating cancer type (Melanoma or RCC). The timing of neoantigen vaccination, administration of subsequent pembrolizumab, optional re-biopsy, disease progression, and patient status (alive or deceased) are shown over a 68-week period.

The primary objective was to evaluate the safety of the intramuscular personalized neoantigen vaccine. Safety was assessed by the incidence and severity of adverse events (AEs), graded according to the National Cancer Institute Common Terminology Criteria for Adverse Events (NCI-CTCAE), version 4.0. Secondary endpoints included immunological outcomes and clinical efficacy, with the latter evaluated by objective response rate (ORR) according to the Immune Response Evaluation Criteria in Solid Tumors (iRECIST), version 1.1.

All study procedures were conducted in accordance with the Declaration of Helsinki and Good Clinical Practice guidelines. The protocol was approved by the Institutional Review Board of the Faculty of Medicine, Chulalongkorn University (COA No. 420/2020, IRB No. 081/62), was registered with the Thai Clinical Trials Registry (TCTR20191003001). Written informed consent was obtained from all participants before enrollment.

### Neoantigen Identification and Peptide Design

#### Sample Collection and Processing

Fresh tumor tissues were collected and snap-frozen. Peripheral blood mononuclear cells (PBMCs) were isolated from blood collected in EDTA tubes and used as the source for germline DNA. Blood samples were centrifuged at 1,600 × g for 15 minutes at 4 °C prior to PBMC isolation using a Ficoll gradient.

#### Genomic and Transcriptomic Sequencing

Genomic DNA was extracted from tumor and PBMC samples using the QIAGEN AllPrep kit. DNA libraries were prepared using MGIEasy FS DNA Library Prep Kit, exomes were captured using SureSelect Human All Exon kits (V6/COSMIC or V7) and sequenced on an MGI DNBSEQ-G400 platform, using a 2 × 150 bp paired-end configuration. The targeted average depth was at least 500x for tumor DNA and 100x for normal DNA. Tumor RNA was isolated, mRNA was enriched by poly-A selection and cDNA libraries for RNA sequencing on the same platform.

#### Bioinformatic Pipeline

Sequencing reads were aligned to the human reference genome (hg38) using Burrows-Wheeler Aligner (BWA) [20] and processed with Genome Analysis Toolkit (GATK) [21]. Somatic variants were identified using a consensus of three variant callers (Mutect2, Strelka, VarScan) [22–24]. HLA class I and II genotypes were determined using HLA-LA [25], ATHLATES [26], and Polysolver [27]. Gene expression was quantified from RNA-seq data using Kallisto [28]. Somatic mutation profile and HLA allele distribution were summarized in Supplementary figure S1 and S2

#### Neoantigen Peptide Design

pVACseq was used to derive and prioritize neoepitopes by integrating somatic variant, HLA-genotype, and gene expression data. MHC binding affinities were predicted using NetMHC, NetMHCII, NetMHCpan, NetMHCIIpan, and MHCFlurry [29–32]. Synthetic long peptides (SLPs) were designed with each spanning 20–35 amino acids in length.

### Vaccine Manufacturing and Formulation

#### Peptide Synthesis

SLPs were manufactured under GMP conditions by commercial vendors (Mimotopes, Australia, GL Biochem, China, and Genscript, China) using solid-phase synthesis and were purified to a minimum of 95% purity. Sterility and safety were confirmed by bioburden and endotoxin testing upon arrival at the clinical site.

#### Vaccine Formulation

In an aseptic isolator, peptides were formulated in 5% dextrose in water (D5W). Peptides were pooled into four groups of 3–5 peptides each. The peptide mixtures were terminally sterilized using a 0.22-micron filter. Immediately before administration, each peptide pool was combined with the adjuvant poly-ICLC (Hiltonol, Oncovir Inc.), a Toll-like receptor 3 (TLR3) agonist. Insoluble peptides were grouped separately into designated pools, specifically Pool A (left arm) and Pool D (right thigh), to prevent them from affecting the filtration or mixing of the soluble peptides. Soluble peptides are alphabetically distributed among the remaining pools.

### Vaccination and Clinical Monitoring

The vaccination regimen consisted of a priming phase and a booster phase. The priming phase included five vaccine doses administered on day 1, 4, 8, 15 and 22, followed by two booster doses at weeks 12 and 20. Each dose contained 0.3 mg of each individual neoantigen peptide combined with 0.5 mg of poly-ICLC. The vaccines were administered intramuscularly at four fixed limb sites. Patients were observed at the clinic for 3 hours following the first dose administration and at least 1 hour in the subsequent doses. Clinical evaluation and adverse event monitoring were performed at each vaccination visit and subsequently every three-month interval during follow-up. Laboratory assessments—including complete blood count, blood urea nitrogen, creatinine, electrolytes, glucose, liver function tests, amylase, lipase, and thyroid-stimulating hormone—were obtained at baseline and at scheduled vaccination and post-vaccination time points. Radiographic evaluations of chest, abdomen, and brain were performed at baseline, week 9, and week 18, and thereafter every three months (Table S1).

### Immunological Monitoring

#### Sample Collection

Blood (40 mL) was collected for immunological analysis at baseline (day 1), during the priming phase (days 8, 15, and 22), and at follow-up visits on weeks 6, 12, 16, 20, and 24. Long-term follow-up assessments were conducted every 3 months thereafter.

#### IFN-γ ELISpot Assay

PBMCs were isolated and stimulated with peptide pools or individual peptides to assess T cell responses. A positive response was defined as a spot count exceeding the mean of negative controls by three standard deviations, with a minimum threshold of 9 Spot-Forming Units (SFU) per 10⁶ cells.

#### Flow Cytometry

Functional profiling of CD4⁺ and CD8⁺ T cells was conducted by measuring intracellular IFN-γ production following stimulation with immunogenic peptides. Phenotypic analysis of T cell memory subsets (e.g., naïve, effector memory, terminally differentiated effector memory was also performed to evaluate changes post-vaccination.

### Tumor Re-Biopsy Analysis

Three patients consented to undergo an optional re-biopsy of a metastatic lesion after receiving vaccine treatment. Tumor tissues collected pre- and post-vaccination were analyzed using multiplex immunohistochemistry (IHC) Vectra system (Akoya Biosciences) to visualize and quantify the density of CD4⁺, CD8⁺, and PD-1⁺ cells.

### Statistical Analysis

In dot distribution plots comparing two subgroups, such as immune responses in ICI-pretreated versus ICI-naïve patients (Figure 2E) or immune cell infiltration before and after treatment measured by cell density (Figure 5), the Mann-Whitney U test was applied. For comparisons involving three or more groups, such as analysis of correlations between peptide characteristic and immunogenicity (Figure S6), the Kruskal Wallis test with Dunn’s multiple-comparison correction was used.

**Figure 2:**
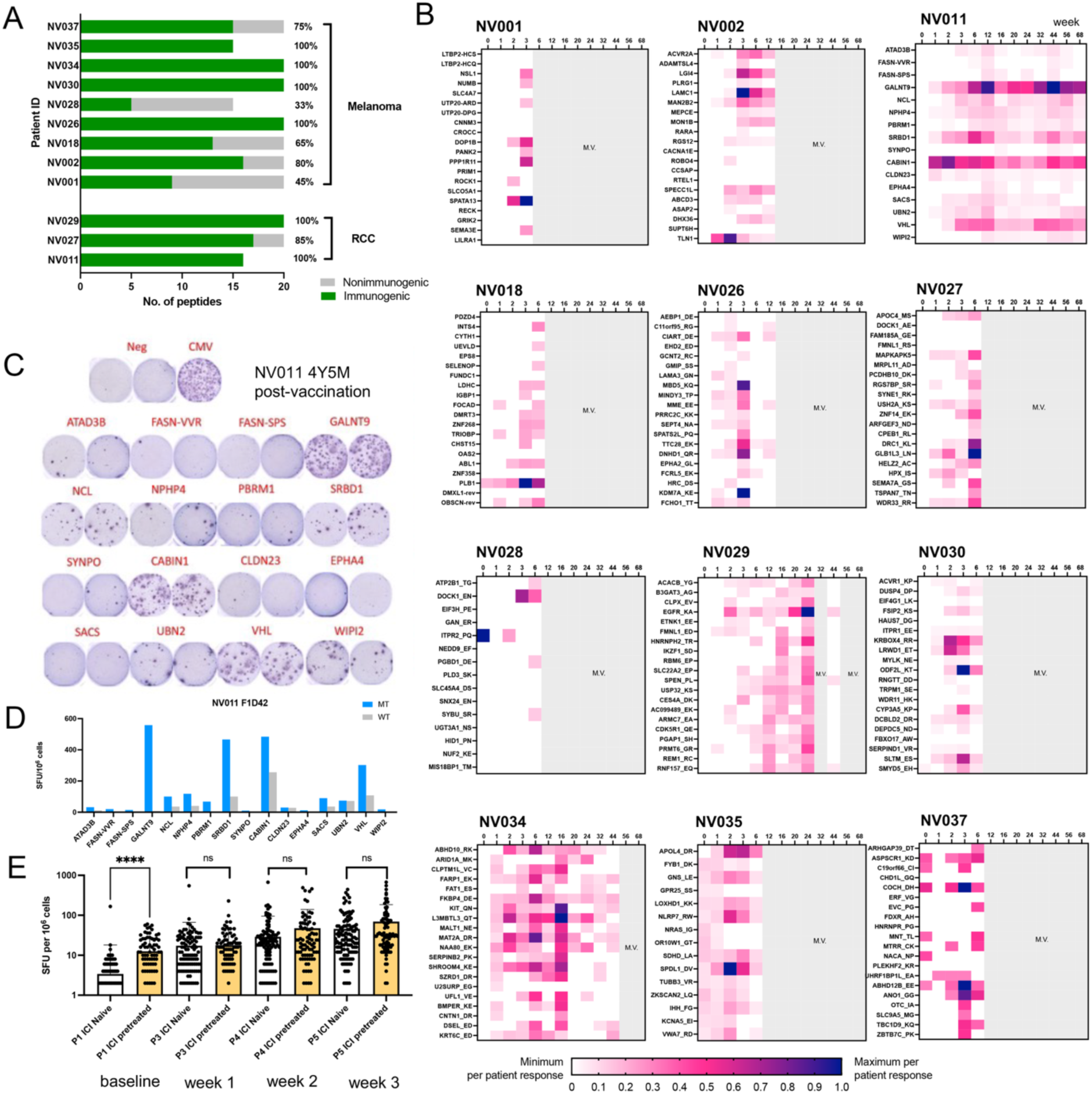
Breadth and kinetics of vaccine-induced neoantigen-specific T-cell responses. (A) A bar chart summarizes the number of immunogenic versus non-immunogenic peptides for each patient, showing that all patients mounted responses to multiple peptides (B) Heatmaps show IFN-γ ELISpot responses for each patient over time against their respective pools of individual neoantigen peptides. The color intensity represents the magnitude of the response, with “m.v.” indicating a missing visit. Responses are detectable as early as week 1 and peak around week 3. (C) The bottom panel shows representative ELISpot wells for patient NV011 at long term response-4 years 5 months after vaccination. (D) Immune responses were neoantigen-specific but not corresponding wild-type counterparts. (E) Comparison of immune checkpoint inhibitor (ICI)-pretreated and ICI-naive groups (**** p from Mann-Whitney U test < 0.0001).

## Results

### Patient Characteristics

A total of 12 eligible patients were enrolled, including 9 melanomas and 3 RCCs. The median age was 56.5 years (range, 48–70). Among the melanoma cohort, 6 patients were female and 3 were male, while all 3 RCC patients were male. None had a history of smoking. At enrollment, 11 of 12 patients (92%) had metastatic disease, while one patient presented with locally recurrent unresectable vulvar melanoma. The median number of prior systemic therapy lines was one. Five patients (42%) had previously received and progressed on immune checkpoint inhibitors (Table 1).

**Table 1:** Patient demographics and clinical characteristics. Summary of baseline characteristics for all 12 patients enrolled in the trial, including age, gender, cancer type, disease stage, and prior history of immune checkpoint inhibitor (ICI) therapy.

| Characteristics | All patients<br>N= 12 (%) |
| --- | --- |
| **Age, median (IQR)** | 56.5 (48-70) |
| **Female, n (%)** | 6 (50%) |
| **ECOG PS, n (%)** |  |
| 0 | 1 (8.3%) |
| 1 | 11 (91.67%) |
| **Comorbidity** |  |
| No | 2 (16.7%) |
| Yes | 10 (83.3%) |
| • Hypertension | 7 (58.3%) |
| • Diabetes | 2 (16.7%) |
| • Dyslipidemia | 5 (41.7%) |
| • Atrial fibrillation | 2 (16.7%) |
| • Chronic hepatitis B infection | 1 (8.3%) |
| • Chronic kidney disease | 1 (8.3%) |
| • Adrenal insufficiency | 1 (8.3%) |
| • Neurofibromatosis | 1 (8.3%) |
| **Cancer type, n (%)** |  |
| Melanoma | 9 (75%) |
| • Acral lentiginous melanoma | 5 (41.7%) |
| • Mucosal melanoma | 4 (33.3%) |
| Renal cell carcinoma (RCC) | 3 (25%) |
| • Clear cell RCC | 2 (16.7%) |
| • Non-clear cell RCC | 1 (8.3%) |
| **Site of metastasis at entry, n (%)** |  |
| Lymph node | 9 (75%) |
| Lung | 7 (58.3%) |
| Skin/Subcutaneous | 6 (50%) |
| Peritoneum | 4 (33.3%) |
| Liver | 3 (25%) |
| Brain | 2 (16.7%) |
| Bone | 2 (16.7%) |
| Pleura | 2 (16.7%) |
| Adrenal gland | 2 (16.7%) |
| Ovary | 1 (8.3%) |
| Bowel | 1 (8.3%) |
| Bladder | 1 (8.3%) |
| **Previous cancer treatment, n (%)** |  |
| Prior surgery | 10 (83.3%) |
| Prior radiation | 6 (50%) |
| Prior systemic treatment for metastatic stage | 9 (75%) |
| • Chemotherapy | 3 (25%) |
| • Targeted therapy | 5 (41.7%) |
| • Immune check point inhibitor | 5 (41.7%) |
| - Nivolumab | 2 (16.7%) |
| - Nivolumab + Ipilimumab | 2 (16.7%) |
| - Pembrolizumab | 1 (8.3%) |
| **Previous line of systemic treatment, n (%)** |  |
| 0 | 3 (25%) |
| 1 | 5 (41.7%) |
| 2 | 3 (25%) |
| 3 | 1 (8.3%) |

### Vaccine Composition and Vaccination

Each patient received a personalized neoantigen vaccine designed from tumor-specific somatic mutations identified by whole-exome sequencing (WES). Most identified somatic variants were missense mutations resulting from single nucleotide variants (SNVs). It is interesting to noted that there was no shared somatic variants detected across the patient cohort, reinforcing the necessity for a personalized approach. Each vaccine consisted of approximately 20 unique synthetic long peptides (SLPs), each 20-35 amino acids in length.

Three patients (NV011, NV028, and NV035) received a reduced number of peptides (16, 15, and 15, respectively) due to a low number of identified mutations or manufacturing constraints. The SLPs were formulated with poly-ICLC, the TLR3 agonist as an adjuvant, and administered intramuscularly.

All patients received five priming doses, but only three (25%) completed the full planned regimen, which included both priming and booster phases (Table S2). Disease progression was the sole reason for vaccine discontinuation.

### Safety and Tolerability

The personalized neoantigen vaccine was safe and well tolerated. All adverse events were grade 1–2, and no serious adverse events, systemic inflammatory responses, or immune-mediated toxicities were observed during the study period. Across all vaccinations, the most common vaccine-related adverse events (VRAEs) were local injection site pain (66.2%) and fever (23.5%). The median onset of VRAEs occurred on the first day of vaccination, with a median duration of 2 days. The frequency of VRAEs was highest at the first priming dose and declined with subsequent doses (Figure 1B, Table S3).

Regarding clinical outcomes, one treatment-naïve patient with clear cell renal cell carcinoma and skin/subcutaneous metastases achieved stable disease for 5.2 months. Disease progression was observed only at subcutaneous sites. The patient’s overall survival exceeds 50 months from vaccine initiation and remains alive at the data cutoff date. The clinical course of all patients, including treatment timelines and outcomes, is depicted in Figure 1C. Among the three patients who completed the full vaccination schedule, all remained evaluable for long-term immune monitoring, with follow-up extending to 68 weeks.

### Vaccine-Induced Systemic Immune Responses

All 12 patients exhibited neoantigen-specific T cell responses to at least one positive neoantigen peptide in their personalized vaccine, as measured by IFN-γ ELISpot assays. On average, 46% of the peptides tested for each patient were immunogenic, with a range of 33% to 100% (Figure 2A). The immune response with IFN-γ responses was detectable as early as one week (Day 8) after the first vaccine dose in most patients (Figure 2B). These responses typically peaked around Day 22 (week 3) or later, following the completion of the priming series (Figure 2B). NV011 demonstrated the longest-surviving paticipant in the cohort, with the measurable immune responses persisting as late as October 2025, corresponding to 4 years and 5 months after the initial vaccination (Figure 2C). Comparative ELISpot analyses confirmed the vaccine’s specificity, demonstrating that T cell responses were significantly more robust against the mutant neoantigen peptides than their corresponding wild-type sequences (Figure 2D). This confirms the vaccine’s capacity to induce mutation-specific immunity without significant off-target reactivity. The immune responses patients previously treated with immune checkpoint inhibitors (ICI-pretreated) were compared with patients who have not received the drug (ICI-naïve) (Figure 2E).The ICI-pretreated group demonstrated significantly higher IFN-γ baseline immune responses, which were measured before the first vaccination dose. At the subsequent visits, however, no significant differences were observed between the two groups. In the ICI-pretreat group, although there is a higher immune response at baseline, we observed a significant increase of immune responses after 21 days in the same manner as the ICI-naïve group.

### Functional Profile and Phenotype of Vaccine-Induced T Cells

To characterize the vaccine-induced T-cell response, flow cytometry was performed on peripheral blood mononuclear cells (PBMCs). The results revealed the induction of IFN-γ-producing CD8⁺ and CD4⁺ T cells upon stimulation with immunogenic peptides. These findings demonstrated that neoantigen vaccination activated both CD8+ and CD4+ T cells (41% CD8+ and 59% CD4+ T cells at the priming phase; 47% CD8+ and 53% CD4+ at the booster phase), without preferentially dominance of either population (Figure 3A). This distribution was consistently observed at the level of individual peptides (Figure 3B and Supplementary Figure S3). Compared with the priming phase, a slight increase in CD8+ proportion was noted at the booster phase (41% versus 47% based on all cases).

**Figure 3:**
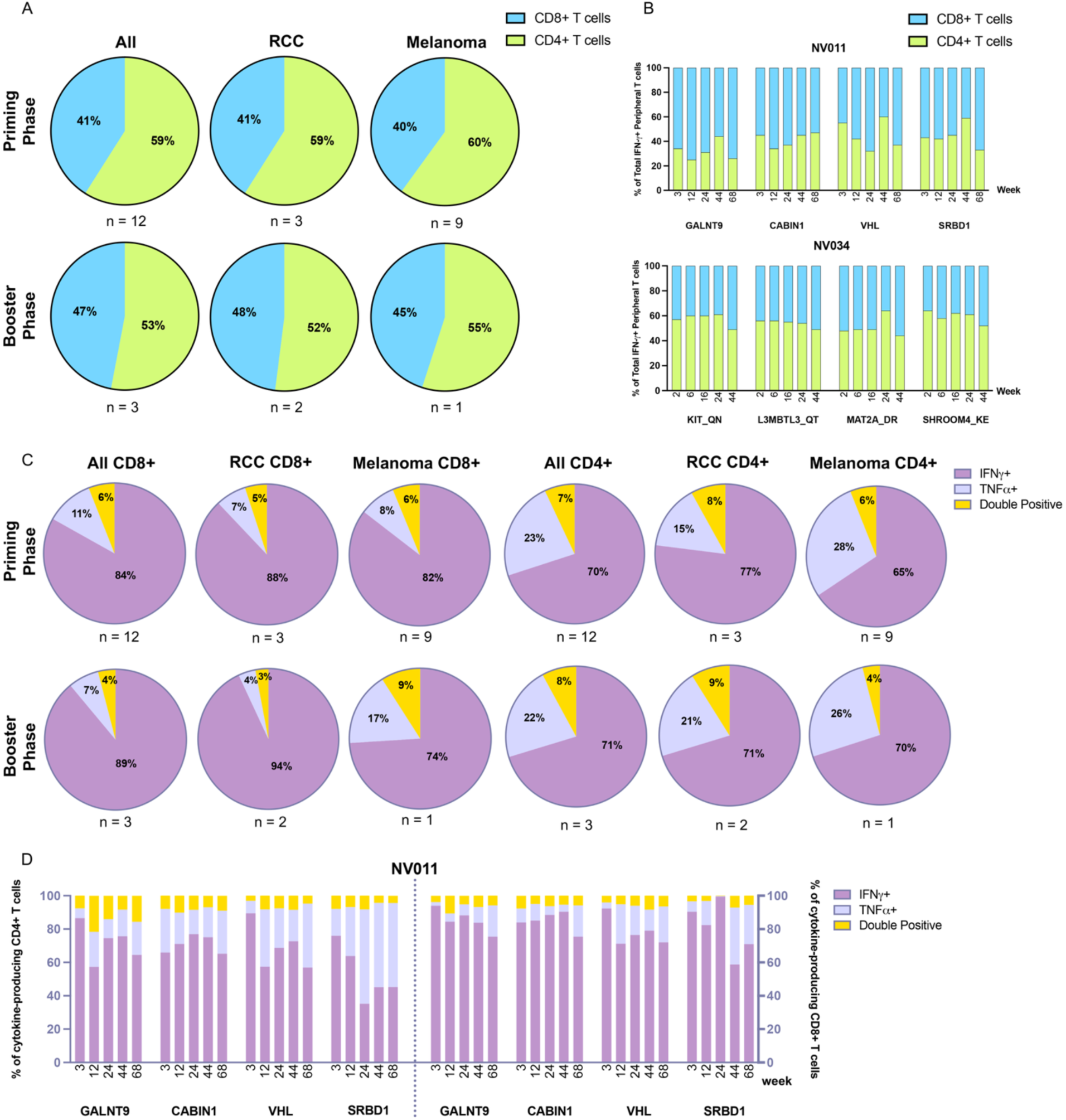
Characterization of neoantigen-specific CD4⁺ and CD8⁺ T-cell responses. (A and B) Pie chart and stacked bar charts showing the relative proportions of IFN-γ-producing CD4⁺ (green) and CD8⁺ (blue) T-cells following stimulation with immunogenic neoantigen peptides. The data demonstrate that the vaccine induced both CD4⁺ and CD8⁺ T-cell responses. (C and D) pie and bar graph showing relative proportion of single positive (IFN-γ or TNF-α) and double positive T cells (IFN-γ and TNF-α).

With respect to pro-inflammatory T cell phenotypes, the majority is IFN-γ-producing T cells. IFN-γ-producing cells were more dominant in CD8+ than CD4+ subsets (84% versus 70% during priming; 89% versus 71% during boosting, based on all cases) (Figure 3C). In contrast, TNF-α-producing T cells were less frequent in CD8+ than CD4+ subsets (11% versus 23% at priming; 7% versus 22% at boosting, based on all patients). The proportion of double-positive cells (IFN-γ and TNF-α) was similar between the two groups (6% versus 7% during priming). These functional profiles were consistent across individual peptides (Figure 3D)

Phenotypic analysis of T cell memory subsets showed dynamic changes following vaccination. Four subpopulations of T cells: Naïve, effector memory (TEM), central memory (TCM) and terminally differentiated effector memory (TEMRA), were examined within CD4+ and CD8+ T cell groups. These were assessed in overall T cells (upper panel) and IFN-γ-positive T cells (lower panel) (Figure 4). In CD4+ subsets, the TEM proportion is higher in IFN-γ-positive T cells, compared with overall T cells, while naïve T cells proportion is nearly absent in IFN-γ-positive T cells. This observation indicates that for neoantigen-specific CD4+ T cells, most naïve cells turned into effector memory. The proportions of TEMRA and TCM varied among patients.

**Figure 4:**
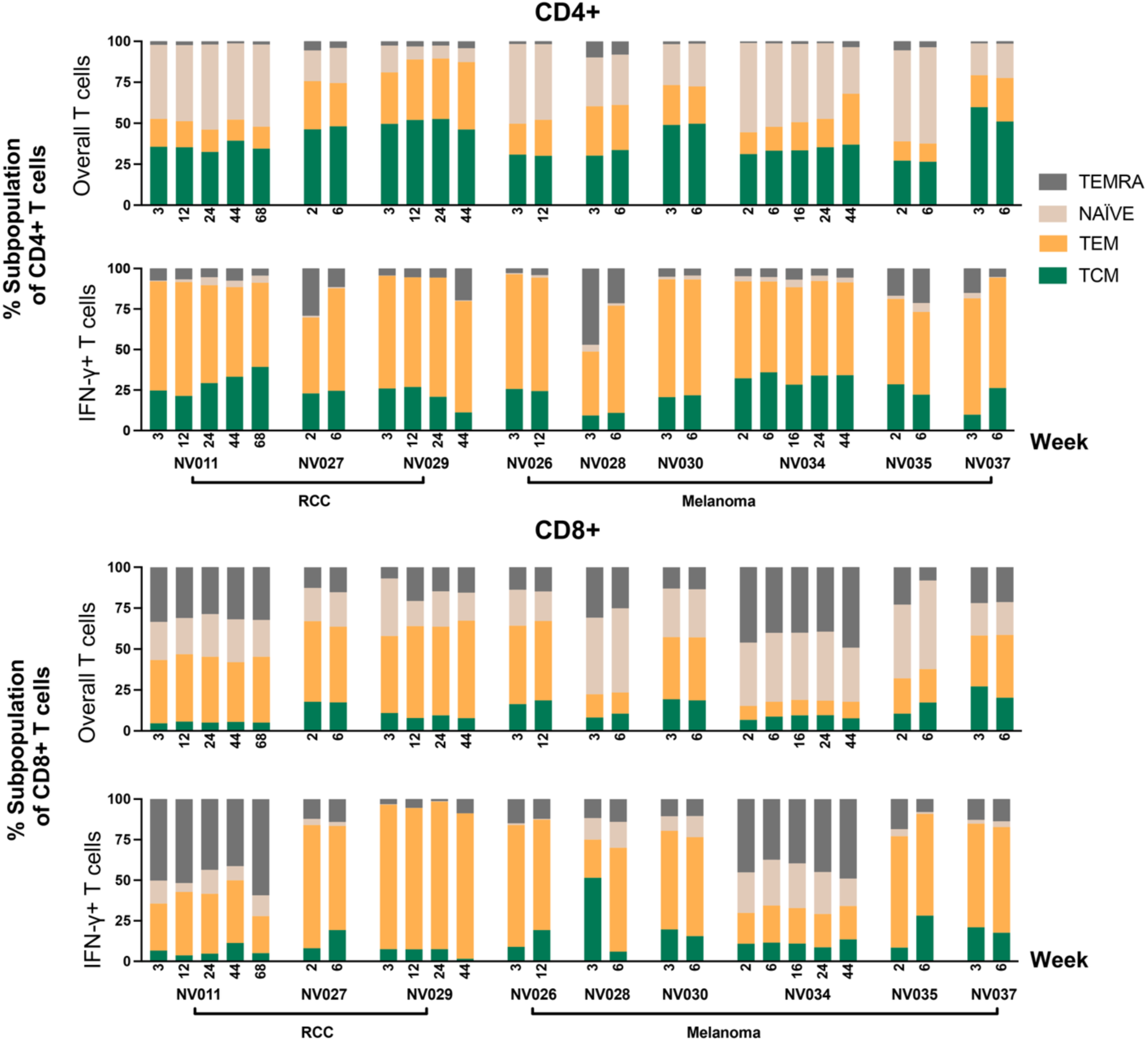
Dynamic changes in T-cell memory subsets following vaccination. Stacked bar charts illustrating the proportional changes in T-cell memory subsets within the CD4⁺ (top) and CD8⁺ (bottom) compartments for each patient over time. The analysis shows a shift from Naïve T-cells (light orange) towards antigen-experienced Effector Memory (TEM, dark orange) for IFN-γ-positive T cells, indicating successful antigen-specific T-cell differentiation.

For CD8+ subsets, patient NV011 and NV034 exhibited higher levels of TEMRA in both overall and IFN-γ-positive T cells. Notably, these patients were the longest survivors in the cohort. Similar to CD4+ T cells, though to a lesser extent, IFN-γ–positive CD8+ T cells showed higher TEM and lower naïve proportions compared with overall CD8+ T cells. This pattern indicates successful differentiation into neoantigen-experienced functional phenotypes in CD8+ T cells.

### Evidence of Immune Remodeling in the Tumor Microenvironment

Three patients (NV011 – RCC, NV026 – melanoma, and NV029 – RCC) consented to an optional re-biopsy of a metastatic lesion after vaccination, allowing for a direct comparison of pathological phenotypes before and after treatment. Tumor tissues collected pre- and post-vaccination were analyzed using multiplex IF to visualize CD4⁺, CD8⁺, and PD-1⁺ cells, along with DAPI nuclear staining. Cell densities were quantified and compared between the two time points (Figure 5). In patient NV011, immunofluorescence analysis revealed a 6.8-fold increase in CD8⁺ T-cell infiltration post-vaccination, suggesting robust immune activation induced by the neoantigen vaccine. Conversely, a reduction in CD4⁺ T-cell density was observed, as confirmed by CD4-specific staining. The density of PD-1⁺ cells also increased 4.3-fold post-vaccination in this patient In the melanoma patient NV026, an increase in CD8⁺ T-cell infiltration and a decrease in CD4⁺ T-cell density were similarly observed following vaccination. A reduction in PD-1⁺ cell density was also noted; however, this was not attributable to pembrolizumab treatment, as the checkpoint inhibitor was administered after the re-biopsy procedure. In patient NV029, no significant changes were detected in CD4⁺ or PD-1⁺ cell densities. While the overall CD8⁺ T-cell density slightly decreased, spatial analysis showed that CD8⁺ cells were more diffusely distributed throughout the tissue post-vaccination, in contrast to the localized clustering seen in pre-vaccination samples.

**Figure 5:**
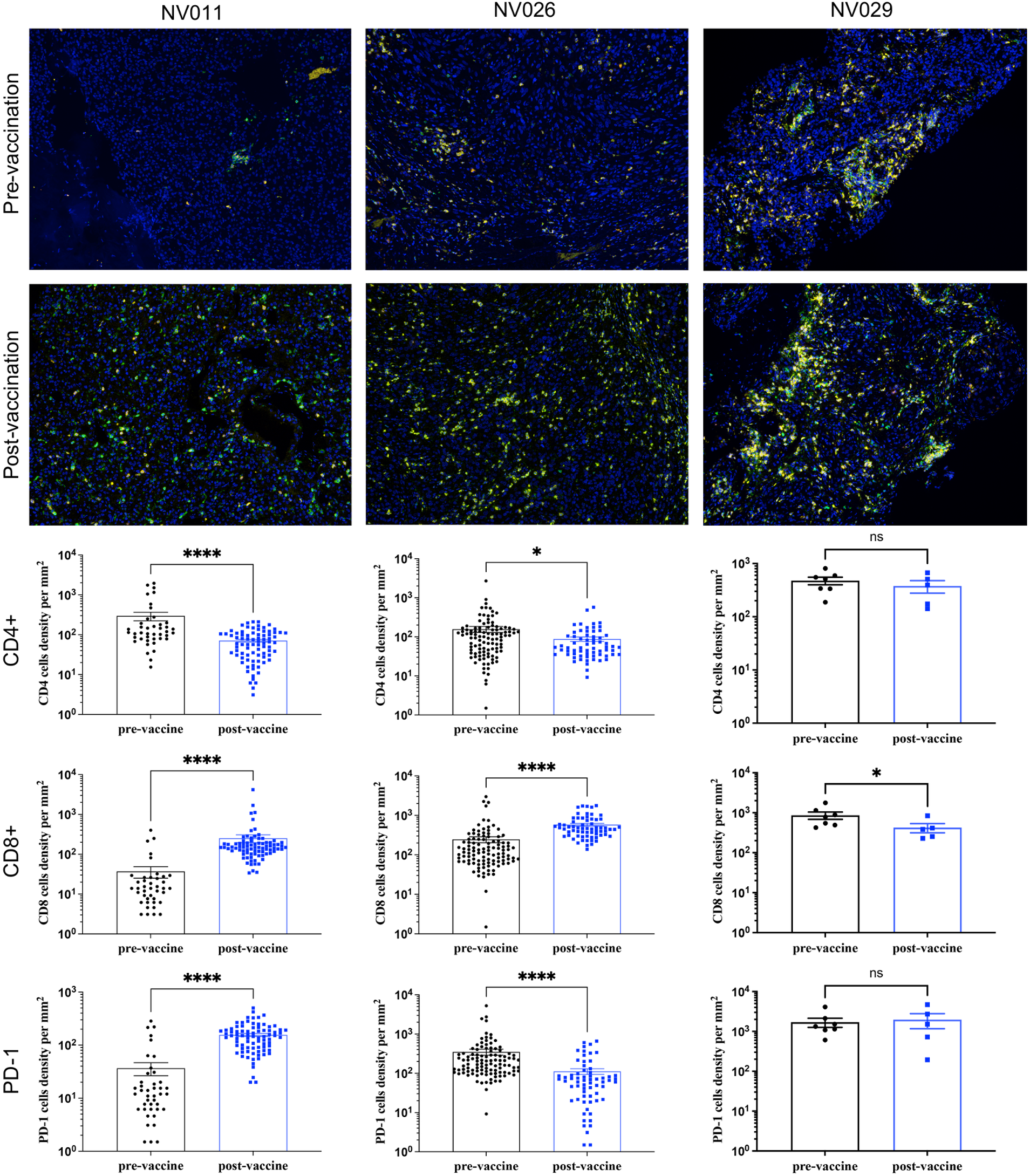
Pathological analysis of the tumor microenvironment. Representative multiplex immunofluorescence images and corresponding quantification of T-cell infiltration in tumor biopsies from patients NV011, NV026, and NV029 taken before and after vaccination. Stains show CD4⁺ T-cells, CD8⁺ T-cells, and PD-1⁺ cells. The charts quantify the change in cell density per mm², revealing a significant increase in CD8⁺ T-cell infiltration post-vaccination in patients NV011 and NV026. Statistical significance is denoted by asterisks (* p < 0.05, **** p < 0.0001 by Mann-Whitney U test).

Overall, re-biopsy provided valuable insights into post-vaccination pathological changes in the tumor microenvironment. Although it was not feasible to collect samples from the exact same lesion, the observable shifts in T-cell infiltration patterns suggest meaningful vaccine-induced immunological remodeling.

### Evidence of Epitope Spreading

Longitudinal immunomonitoring of T cell responses provided an opportunity to assess for epitope spreading, a phenomenon in which a therapeutic intervention induces an immune response that expands to recognize additional antigens not included in the original neoantigen vaccine. The epitope spreading experiments were performed in two patients (NV011 and NV034). Clear evidence of epitope spreading was observed in patient NV011, in whom T cell responses against 8 mutated SLPs not included in the original neoantigen vaccine were detected (Supplementary Figure S4). These newly detectable responses to non-immunized neoantigen peptides emerged primarily during the boosting phase of the trial (after week 12). These responses were minimal at baseline and during the early post-priming period, indicating a dynamic and broadening of the T cell repertoire over time. This finding suggests that the initial vaccine-driven T cell response may have led to sufficient tumor cell lysis to release new tumor-specific and tumor-associated antigens, facilitating an expanded, endogenous anti-tumor immune response against targets beyond those in the vaccine itself. In contrast, strong evidence of epitope spreading was not observed in patient NV034, in whom 12 non-immunized neoantigen peptides were tested.

### Analysis of Factors Influencing Immunogenicity

To identify determinants of immunogenicity, we analyzed 239 SLPs from all 12 patients against multiple variables including peptide characteristics and genomic features. Analysis of peptide properties revealed several key factors (Supplementary Figure S5). First, with respect to MHC binding predictions, SLPs containing predicted epitopes for both MHC class I and class II molecules were significantly more immunogenic than those with predictions for class I alone. However, neither the strong predicted binding affinity nor the % rank correlated with immunogenicity.

Second, among amino acid properties, mutations involving a proline residue were associated with significantly reduced immunogenicity, possibly due to structural disruption. In contrast, substitutions involving charged or hydrophobic bulky residues did not exert a significant impact. Cysteine-containing peptides within the epitope regions exhibit stronger immune response than peptides without cysteine. Other physicochemical properties such as overall peptide solubility were also not found to be major determinants of the immune response. Furthermore, analysis of ELISpot responses across the four different injection pools revealed no significant differences, indicating that the injection site did not influence immunogenicity in this study.

Analysis of genomic features showed that neoantigens derived from amino acid insertions or deletions (in-frame indels) were significantly more immunogenic than those from missense mutations. Lastly, somatic variants with a high DNA variant allele frequency (VAF) were associated with lower immunogenicity. These observations require validations in a larger dataset. Gene expression levels of the source protein did not correlate with the immunogenicity of the resulting peptide.

## Discussion

This IMPACT Phase Ib trial of personalized neoantigen peptide vaccine administered intramuscular with poly-ICLC demonstrates that it is safe, feasible, and highly immunogenic in patients with advanced melanoma or RCC, irrespective of prior ICI exposure. Notably, the vaccine elicited robust, de novo, mutation-specific both CD8+ and CD4+ T cell responses in all 12 patients, including those who were heavily pretreated. The safety profile was well tolerated, with only grade 1-2 vaccine-related side effects without immune-mediated events. To our knowledge, this is the first trial to report the safety of personalized cancer neoantigen peptide-based vaccines delivered solely via the intramuscular route.

Detailed immune profiling at multiple early timepoints provided significant insight into the kinetics of the vaccine-induced response. IFN-γ ELISpot responses were observed as early as one to two weeks post-vaccination, peaking around weeks 3-6. Specificity was confirmed by comparing responses through mutant against wild-type peptide comparisons, reinforcing the precision of the personalized vaccine design. Furthermore, two longest-surviving participants (NV011 and NV034) mounted early and robust immune responses, suggesting a potential, preliminary link between the quality of the immune response and clinical outcome that needed a further investigation. Our study was able to compare patients who were previously treated with immune checkpoint inhibitors and the naive group. Although we observed the high immune response at baseline in the ICI-pretreat group (N = 5), the ICI-pretreat immunity can still be boosted to achieve positive immune response after completing the five priming doses. There is no significant difference in immune response between ICI-pretreated and ICI-naïve group after priming vaccination started.

A noteworthy finding was the induction of robust CD8⁺ T-cell responses associated with the intramuscular vaccine administration. Immunomonitoring revealed the robust *de novo* induction both CD8+ and CD4+ T-cell responses at priming and booster phases, in both melanoma and RCC patients. This data confirmed the results from preclinical studies [19]. The IM route was not only safe but also a simple way to achieve CD8+ T cell responses. This contrasts with several other neoantigen vaccine trials that have primarily reported strong, but often CD4⁺-dominant, T-cell responses. We hypothesize that the intramuscular route of administration, combined with the use of Poly-ICLC, may have contributed to this favorable CD8⁺ T-cell polarization. While intradermal or subcutaneous injections target the skin’s dense network of antigen-presenting cells (APCs) known to be efficient at CD4⁺ T-cell activation, IM injection into muscle tissue creates a different immunological environment. The local inflammation caused by the injection, potentiated by a strong Type-1 polarizing adjuvant like the TLR3-agonist Poly-ICLC, can promote the recruitment of dendritic cells that are highly capable of cross-presentation—a critical pathway for priming cytotoxic CD8⁺ T-cells [33, 34]. This observation is hypothesis-generating and suggests that the route of administration is a key variable that can be optimized to elicit the desired balance of helper and cytotoxic T-cell responses in cancer vaccination. Overall, the data indicates that IM administration is a viable alternative with practical advantages for stimulating CD8+ T cell, along with CD4+ ones. With the simplicity of intramuscular injection, it is suitable for clinical applications.

A pivotal observation was that the two patients who achieved durable disease stabilization and long-term survival, NV011 and NV034, showed a significant enrichment of neoantigen-specific T cells with a TEMRA phenotype. While a high proportion of terminally differentiated cells might seem counterintuitive for a durable response, this is an increasingly expected hallmark of an effective, mature anti-tumor response in the context of chronic cancer settings [35, 36]. This indicates that the vaccine successfully remodeled the immune compartment to establish a state of persistent and highly functional cytotoxic activity. Furthermore, the predominance of this chronically stimulated T-cell phenotype has direct therapeutic implications. The same process that drives the development of these potent TEMRA cells also puts them at high risk of T-cell exhaustion, which is often mediated through inhibitory pathways like PD-1. This creates a compelling rationale for combining the vaccine with a checkpoint inhibitor. The vaccine serves to build and expand the army of tumor-specific T-cells, while an anti-PD-1/PD-L1 antibody would ensure this army remains functional within the tumor microenvironment [37]. Such a strategy holds the potential to sustain the anti-tumor response and achieve even more durable clinical outcomes.

The immunopathological changes observed in this study provide a strong rationale for future combination therapies. Among three patients who consented to re-biopsy, we observed a significant increase in CD8⁺ T cell infiltration into the tumor microenvironment post-vaccination in two patients. This demonstrates the vaccine’s ability not only to stimulate CD8+ T cell response but to drive functional T cells traffic to the tumor site. Crucially, in patient NV011, this was accompanied by a 4.3-fold increase in PD-1⁺ cells, suggesting that while T cells were trafficking to the tumor, they may have been subsequently suppressed by exhaustion pathways within the tumor microenvironment. This exhaustion driven data correlates with the high TEMRA proportion observed in NV011 but not in other RCC patients. This dual finding strongly supports a strategy of combining this vaccine with an anti-PD-1/PD-L1 checkpoint inhibitor. The vaccine could act to “prime” the tumor by increasing T cell infiltration, creating a more favorable environment for an ICI to then “unleash” the anti-tumor response. This is further supported by the detection of epitope spreading in patient NV011, which signifies a broadening of the anti-tumor response beyond the original vaccine targets. Overall the observations are the hallmark of an effective CD8+ and CD4+ immune response that drive therapeutically antitumor killing.

An analysis of determinants to immunogenicity revealed key findings to optimize peptide-based neoantigen design. The TESLA cohort has been pioneered to identify major predictors to improve immunogenicity [38]. And our clinical dataset further supports and expands these findings. Peptides containing both MHC class I and II predicted epitopes elicited better immunogenicity, emphasizing the importance of engaging CD8+ and CD4+ responses. In contrast, predicted binding affinity and percentile rank did not correlate with immunogenicity, suggesting the limitation of current algorithm, the need for improved TCR-binding prediction, or the presence of additional determinants of antigen presentation that remain uncharacterized.

Amino acid alterations are also influenced immunogenicity. Proline substitution strongly reduced immune responses, likely due to structural disruption, whereas cysteine-containing peptides enhanced immunogenicity. This suggested that cysteine should not be excluded from SLP selection despite concerns about disulfide-related side products during manufacturing. Other biochemical features, such as changes in bulkiness, charges, or solubility, were not major contributors to immunogenicity. Consistently, injection site assignment and peptide solubility showed no impact on immune response.

From a genomic perspective, in-frame indel-derived mutations were significantly more immunogenic, consistent with prior reports [39]. Low variant allele frequency was associated with increased immunogenicity, highlighting the relevance of subclonal mutations and supporting the inclusion of both clonal and subcloncal mutations to broaden the immune repertoire. In contrast to previous studies [11, 38], our gene expression levels did not correlate with peptide immunogenicity, indicating that transcriptional abundance of the source protein may not directly predict T cell recognition.

Taken together, these findings enable the multifactorial nature of neoantigen-stimulated immunogenicity and provide practical guidance for SLP design. Integrating peptide-level features (dual MHC binding, amino acid composition) with genomic attributes (in-frame indel origin, variant allele frequency) may improve the neoantigen prioritization and enhance antitumor responses. Larger datasets will be needed to validate these features and further refine personalized neoantigen vaccine design strategies.

The limitations of this study include the small sample size and limited long-term follow-up in heavily pretreated metastatic patients who had progressive disease and no effective subsequent treatment options. Although strong immunogenic responses were achieved, no objective clinical response was not observed. Importantly, because the primary endpoints of this Phase I study were to evaluate safety and assess immunogenic signals, the study was not designed or powered to determine clinical efficacy, especially in the absence of combination therapy with ICIs. Nevertheless, our findings provide a rationale for future trials to investigate intramuscular SLP vaccination in combination with ICIs earlier in the treatment course to enhance clinical benefit. Additional strategies, such as extended peptide boosting regimens or alternative adjuvants, should also be evaluated to improve the durability of immune responses.

In conclusion, this study demonstrates that a personalized intramuscular neoantigen SLP vaccine can safely elicit rapid, robust, and mutation-specific both CD8+ and CD4+ T-cell responses in patients with advanced cancer, even in the absence of concurrent checkpoint blockade. These results provide a compelling immunological rationale for advancing this approach into future combination strategies with ICIs to maximize clinical benefit.

## Authors’ Contributions

None.

## Data Availability

All data in this study are available upon request to the authors

## Acknowledgments

This study is supported by Donation from Thai people, Program Management Unit for Competitiveness (PMUC) grant 2563 and 2564, Ratchadapiseksompotch Fund, Faculty of Medicine, Chulalongkorn University, grant number RA-MF-45/64, Ratchadapiseksompotch Matching Fund from Faculty of Medicine, Chulalongkorn University, and Research University Network Thailand. We appreciate Sumanee Nilgate for assisting us in testing the sterility of the vaccine. We also thank the Medical Oncology Unit at King Chulalongkorn Memorial Hospital, Bangkok, Thailand, for their support of this study.

## Reference

1. Jenkins, R.W., D.A. Barbie, and K.T. Flaherty, Mechanisms of resistance to immune checkpoint inhibitors. British Journal of Cancer, 2018. 118(1): p. 9–16.

2. Ribas, A. and J.D. Wolchok, Cancer immunotherapy using checkpoint blockade. Science, 2018. 359(6382): p. 1350-1355.

3. Blass, E. and P.A. Ott, Advances in the development of personalized neoantigen-based therapeutic cancer vaccines. Nature Reviews Clinical Oncology, 2021. 18(4): p. 215–229.

4. Schumacher, T.N. and R.D. Schreiber, Neoantigens in cancer immunotherapy. Science, 2015. 348(6230): p. 69-74.

5. Weber, J.S., et al., Individualised neoantigen therapy mRNA-4157 (V940) plus pembrolizumab versus pembrolizumab monotherapy in resected melanoma (KEYNOTE-942): a randomised, phase 2b study. The Lancet, 2024. 403(10427): p. 632–644.

6. Sahin, U., et al., Personalized RNA mutanome vaccines mobilize poly-specific therapeutic immunity against cancer. Nature, 2017. 547(7662): p. 222-226.

7. Zhang, X., et al., Neoantigen DNA vaccines are safe, feasible, and induce neoantigen-specific immune responses in triple-negative breast cancer patients. Genome Medicine, 2024. 16(1): p. 131.

8. Ding, Z., et al., Personalized neoantigen pulsed dendritic cell vaccine for advanced lung cancer. Signal Transduction and Targeted Therapy, 2021. 6(1): p. 26.

9. Guo, Z., et al., Durable complete response to neoantigen-loaded dendritic-cell vaccine following anti-PD-1 therapy in metastatic gastric cancer. npj Precision Oncology, 2022. 6(1): p. 34.

10. Ingels, J., et al., Neoantigen-targeted dendritic cell vaccination in lung cancer patients induces long-lived T cells exhibiting the full diVerentiation spectrum. Cell Reports Medicine, 2024. 5(5): p. 101516.

11. Ott, P.A., et al., An immunogenic personal neoantigen vaccine for patients with melanoma. Nature, 2017. 547(7662): p. 217–221.

12. Braun, D.A., et al., A neoantigen vaccine generates antitumour immunity in renal cell carcinoma. Nature, 2025. 639(8054): p. 474–482.

13. Zhang, Z., et al., Neoantigen: A New Breakthrough in Tumor Immunotherapy. Frontiers in Immunology, 2021. Volume 12 - 2021.

14. Wu, D.-W., et al., Personalized neoantigen cancer vaccines: current progression, challenges and a bright future. Clinical and Experimental Medicine, 2024. 24(1): p. 229.

15. Xie, N., et al., Neoantigens: promising targets for cancer therapy. Signal Transduction and Targeted Therapy, 2023. 8(1): p. 9.

16. Hu, Z., et al., Personal neoantigen vaccines induce persistent memory T cell responses and epitope spreading in patients with melanoma. Nature Medicine, 2021. 27(3): p. 515–525.

17. Sultan, H., A.M. Salazar, and E. Celis, Poly-ICLC, a multi-functional immune modulator for treating cancer. Seminars in Immunology, 2020. 49: p. 101414.

18. Blass, E., et al., A multi-adjuvant personal neoantigen vaccine generates potent immunity in melanoma. Cell, 2025. 188(19): p. 5125–5141.e27.

19. Sultan, H., et al., The route of administration dictates the immunogenicity of peptide-based cancer vaccines in mice. Cancer Immunology, Immunotherapy, 2019. 68(3): p. 455–466.

20. Li, H. and R. Durbin, Fast and accurate short read alignment with Burrows–Wheeler transform. Bioinformatics, 2009. 25(14): p. 1754–1760.

21. McKenna, A., et al., The Genome Analysis Toolkit: A MapReduce framework for analyzing next-generation DNA sequencing data. Genome Research, 2010. 20(9): p. 1297–1303.

22. Cibulskis, K., et al., Sensitive detection of somatic point mutations in impure and heterogeneous cancer samples. Nature Biotechnology, 2013. 31(3): p. 213–219.

23. Saunders, C.T., et al., Strelka: accurate somatic small-variant calling from sequenced tumor–normal sample pairs. Bioinformatics, 2012. 28(14): p. 1811–1817.

24. Koboldt, D.C., et al., VarScan 2: Somatic mutation and copy number alteration discovery in cancer by exome sequencing. Genome Research, 2012. 22(3): p. 568–576.

25. Dilthey, A.T., et al., HLA*LA—HLA typing from linearly projected graph alignments. Bioinformatics, 2019. 35(21): p. 4394–4396.

26. Liu, C., et al., ATHLATES: accurate typing of human leukocyte antigen through exome sequencing. Nucleic Acids Research, 2013. 41(14): p. e142–e142.

27. Shukla, S.A., et al., Comprehensive analysis of cancer-associated somatic mutations in class I HLA genes. Nature Biotechnology, 2015. 33(11): p. 1152–1158.

28. Bray, N.L., et al., Near-optimal probabilistic RNA-seq quantification. Nature Biotechnology, 2016. 34(5): p. 525–527.

29. O’Donnell, T.J., A. Rubinsteyn, and U. Laserson, MHCflurry 2.0: Improved Pan-Allele Prediction of MHC Class I-Presented Peptides by Incorporating Antigen Processing. Cell Systems, 2020. 11(1): p. 42–48.e7.

30. Andreatta, M. and M. Nielsen, Gapped sequence alignment using artificial neural networks: application to the MHC class I system. Bioinformatics, 2016. 32(4): p. 511–517.

31. Jensen, K.K., et al., Improved methods for predicting peptide binding aVinity to MHC class II molecules. Immunology, 2018. 154(3): p. 394–406.

32. Nilsson, J.B., et al., Accurate prediction of HLA class II antigen presentation across all loci using tailored data acquisition and refined machine learning. Science Advances. 9(47): p. eadj6367.

33. Embgenbroich, M. and S. Burgdorf, Current Concepts of Antigen Cross-Presentation. Frontiers in Immunology, 2018. Volume 9 - 2018.

34. MacNabb, B.W., et al., Dendritic cells can prime anti-tumor CD8+ T cell responses through major histocompatibility complex cross-dressing. Immunity, 2022. 55(6): p. 982–997.e8.

35. Blank, C.U., et al., Defining ‘T cell exhaustion’. Nature Reviews Immunology, 2019. 19(11): p. 665–674.

36. Gattinoni, L., et al., T memory stem cells in health and disease. Nature Medicine, 2017. 23(1): p. 18–27.

37. Hu, Z., P.A. Ott, and C.J. Wu, Towards personalized, tumour-specific, therapeutic vaccines for cancer. Nature Reviews Immunology, 2018. 18(3): p. 168–182.

38. Wells, D.K., et al., Key Parameters of Tumor Epitope Immunogenicity Revealed Through a Consortium Approach Improve Neoantigen Prediction. Cell, 2020. 183(3): p. 818–834.e13.

39. Turajlic, S., et al., Insertion-and-deletion-derived tumour-specific neoantigens and the immunogenic phenotype: a pan-cancer analysis. The Lancet Oncology, 2017. 18(8): p. 1009–1021.

