## Supplementary Figures and Tables for "IMPACT - Phase Ib Trial of Intramuscular Personalized Neoantigen Synthetic Long Peptide Vaccines in Patients with Advanced Melanoma and Renal Cell Carcinoma"

### Supplementary Tables

#### Supplementary Table S1. Schedule of vaccination, clinical assessments, and laboratory monitoring

| | Treatment period | | | | | | | | | | | | Follow up Period Follow up (+/-7 Days) | | | |
|---|---|---|---|---|---|---|---|---|---|---|---|---|---|---|---|---|
| | Priming phase | | | | | | | Booster phase | | | | | | | | |
| Visit Name | P1 | P2 | P3 | P4 | P5 | F1 | I1 | B1 | F2 | I2 | B2 | F3 | V1 | V2 | V3 | V4 |
| Timepoint | D 1 | D 4 | D 8 | D 15 | D 22 | W 6 | W 9 | W 12 | W 16 | W 18 | W 20 | W 24 | W 12 | W 24 | W 36 | W 48 |
| Concomitant medication, procedures | X | X | X | X | X | X | X | X | X | X | X | | X | X | X | X |
| Vital signs[a] | X | X | X | X | X | X | X | X | X | X | X | | X | X | X | X |
| ECOG performance status | X | X | X | X | X | X | X | X | X | X | X | | X | X | X | X |
| Physical examination[b] | X | X | X | X | X | X | X | X | X | X | X | | X | X | X | X |
| Chest and Whole abdomen (CT/MRI)[c] | | | | | | | X | | | X | | | X | X | X | X |
| Brain (CT/MRI)[c] | | | | | | | X | | | X | | | X | X | X | X |
| Response assessment | | | | | | | X | | | X | | | X | X | X | X |
| EKG 12 leads | X | | | | | | | | | | | | | | | |
| Haematology | X | | X | X | X | X | X | X | X | X | X | | X | X | X | X |
| Serum chemistry | X | | X | X | X | X | X | X | X | X | X | | X | X | X | X |
| Thyroid tests | | | | | | | | | | X | | | X | | X | |
| Blood for PBMC | X | | X | | X | X | | X | | | X | | X | | | |
| Urine analysis | X | | X | X | X | X | X | X | X | X | X | | X | X | X | X |
| Neoantigen peptide vaccine administration | X | X | X | X | X | | | X | | | X | | | | | |
| Adverse Events | X | X | X | X | X | X | X | X | X | X | X | | X | X | X | X |

Abbreviations: P: Priming; F: Follow-up; I: Imaging; V: Visit; D: Day; W: Week; ECOG: Eastern Cooperative Oncology Group; CT: computed tomography scan; MRI: Magnetic Resonance Imaging; PBMC: Peripheral blood mononuclear cells

a. Vital signs include body temperature, pulse rate and systolic and diastolic blood pressure. Vital signs will be assessed prior to the administration of each dose of vaccine then every 15 minutes until 60 minutes. At the first dose of vaccine (P1), vital signs will be assessed at 2 and 4 hours after the administration of vaccine also.

b. Physical examination will be assessed prior to the administration of each dose of vaccine in priming and booster phase. Except height will only be measured at screening visit. Weight will be measured at screening visit and prior to the administration of each dose of vaccine.

c. Radiologic assessment should be the same modality throughout the period of the study.

**Supplementary Table S2: Cancer Vaccination**

| Cancer vaccination | All patients N= 12 (%) |
|---|---|
| **Priming phase, n (%)** | |
| P1 | 12 (100%) |
| P2 | 12 (100%) |
| P3 | 12 (100%) |
| P4 | 12 (100%) |
| P5 | 12 (100%) |
| **Booster phase, n (%)** | |
| B1 | 5 (41.7%) |
| B2 | 3 (25%) |

**Supplementary Table S3: Incidence and severity of vaccine related adverse events (VRAEs)**

| VRAE | P1 N= 12 (%) | P2 N= 12 (%) | P3 N= 12 (%) | P4 N= 12 (%) | P5 N= 12 (%) | B1 N= 5 (%) | B2 N= 3 (%) |
|---|---|---|---|---|---|---|---|
| **Any VRAE, N (%)** | 11 (91.7%) | 10 (83.3%) | 10 (83.3%) | 10 (83.3%) | 4 (33.3%) | 1 (20%) | 0 |
| Grade 1 | 7 (58.3%) | 8 (66.7%) | 6 (50%) | 8 (66.7%) | 4 (33.3%) | 1 (20%) | |
| Grade 2 | 4 (33.3%) | 2 (16.7%) | 4 (33.3%) | 2 (16.7%) | 0 | 0 | |
| **Local Pain** | 11 (91.7%) | 10 (83.3%) | 9 (75%) | 10 (83.3%) | 4 (33.3%) | 1 (20%) | |
| Grade 1 | 7 (58.3%) | 8 (66.7%) | 6 (50%) | 9 (75%) | 4 (33.3%) | 1 (20%) | |
| Grade 2 | 4 (33.3%) | 2 (16.7%) | 3 (25%) | 1 (8.3%) | 0 | | |
| **Fever** | 5 (41.7%) | 2 (16.7%) | 1 (8.3%) | 3 (25%) | 4 (33.3%) | 1 (20%) | |
| Grade 1 | 5 (41.7%) | 2 (16.7%) | 1 (8.3%) | 3 (25%) | 4 (33.3%) | 1 (20%) | |
| **Myalgia** | 1 (8.3%) | 1 (8.3%) | 0 | 2 (16.7%) | 0 | 1 (20%) | |
| Grade 1 | 1 (8.3%) | 1 (8.3%) | 0 | 1 (8.3%) | 0 | 1 (20%) | |
| Grade 2 | 0 | 0 | 0 | 1 (8.3%) | 0 | 0 | |
| **Nausea/Vomiting** | 0 | 1 (8.3%) | 2 (16.7%) | 0 | 0 | 0 | |
| Grade 1 | 0 | 1 (8.3%) | 1 (8.3%) | 0 | 0 | 0 | |
| Grade 2 | 0 | 0 | 1 (8.3%) | 0 | 0 | 0 | |

### Supplementary Table S4: Representativeness of Study Participants

| | |
|---|---|
| Cancer type(s)/ stage(s)/condition | Metastatic or unresectable melanoma and renal cell carcinoma (RCC) |
| Sex | Melanoma in Thailand occurs in both males and females, whereas RCC is more common in men, with global epidemiologic data showing a male-to-female ratio of approximately 2:1.<br>In our study, the sex distribution differed between the two cancer types enrolled. Among the nine melanoma patients, six were female and three were male, while all three RCC patients were male. Overall, this pattern generally aligns with expected sex trends for each disease: melanoma shows a relatively balanced sex distribution, whereas RCC remains predominantly male.<br>However, given the small overall sample size (n = 12), the combined sex distribution may not fully represent the broader Thai population with metastatic melanoma or RCC, especially in settings where referral patterns and access to immunotherapy vary. |
| Age | Melanoma and RCC in Thailand are most commonly diagnosed in middle-aged to older adults, with reported median ages in the mid-50s to early 60s. The median age of participants in our trial was **56.5 years (IQR 48–70)**, which is consistent with the expected age range of patients with metastatic melanoma or RCC treated at tertiary centers. |
| Race/Ethnicity | All participants in this study were ethnically Thai (Asian).<br>The vast majority of melanoma and RCC cases in Thailand occur in ethnically Thai or other Southeast Asian populations.<br>Although the enrolled population is representative of the local racial/ethnic distribution, it does not reflect the broader global variability in melanoma or RCC, especially considering known racial differences in melanoma incidence and RCC subtype distribution in Western populations. |
| Geography | All participants were enrolled at King Chulalongkorn Memorial Hospital, a national tertiary-care academic center located in Bangkok, Thailand.<br>Most patients with metastatic melanoma or RCC who require specialized care in Thailand are referred to large urban tertiary centers. However, relying on a single-center metropolitan population may underrepresent patients from rural provinces, where healthcare access, referral patterns, and disease presentation may differ. |
| Other considerations | Patients were required to have ECOG 0–1 and were excluded if they had active autoimmune disease or active infection, which is standard for early-phase immunotherapy trials but may exclude real-world patients with comorbidities.<br>Enrollment included patients who had progressed on or had no access to standard systemic therapies. Access to immune checkpoint inhibitors (ICIs) in Thailand remains limited due to cost and insurance restrictions.<br>Consequently, patients eligible for and treated at tertiary centers may differ from the broader national population in terms of socioeconomic status and treatment availability.<br>Prior ICI exposure was allowed, which reflects contemporary treatment patterns but may not represent patients who receive non-ICI-based care due to availability constraints. |

### Supplementary Figures

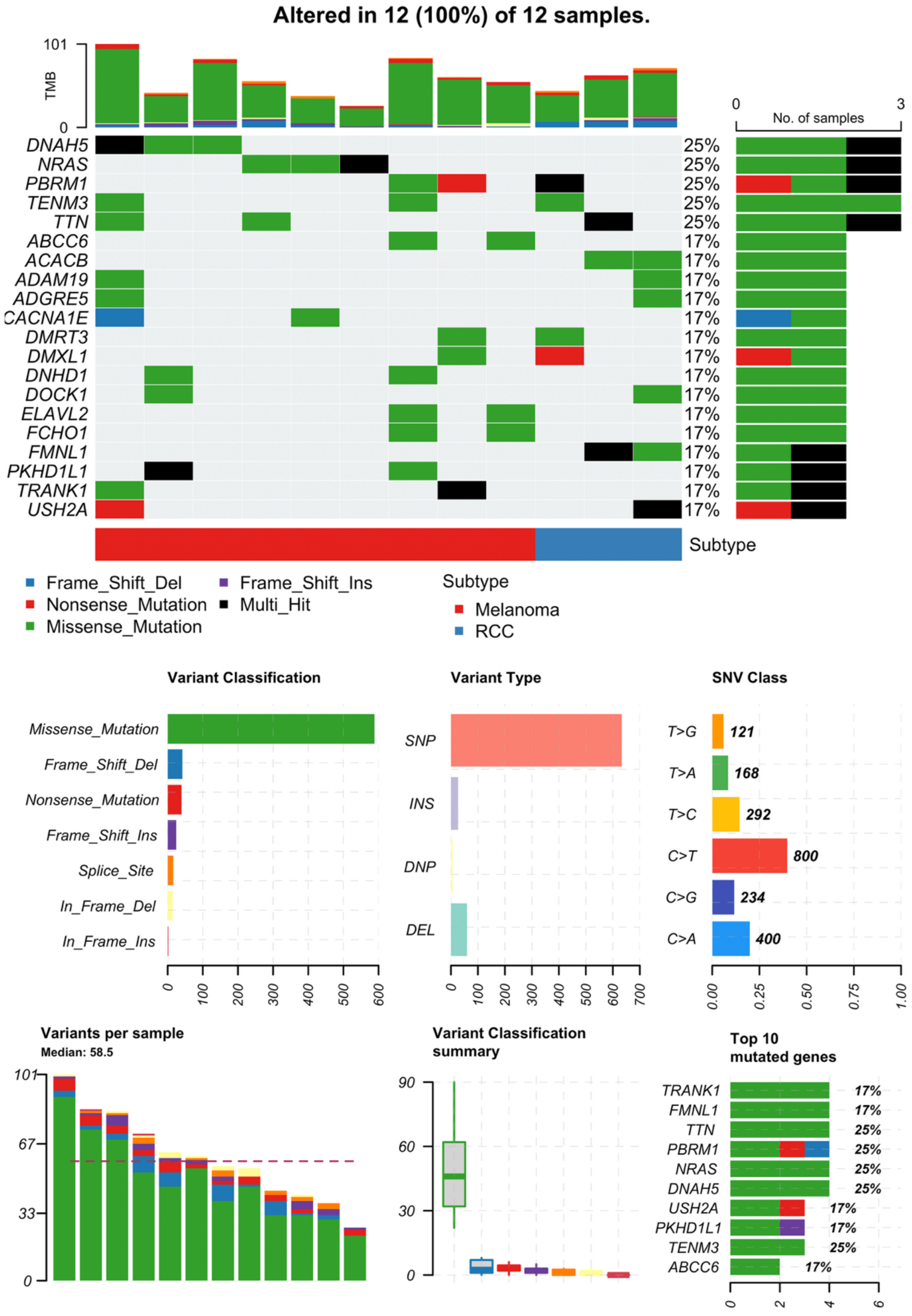


Supplementary Figure S1: Somatic mutation profile

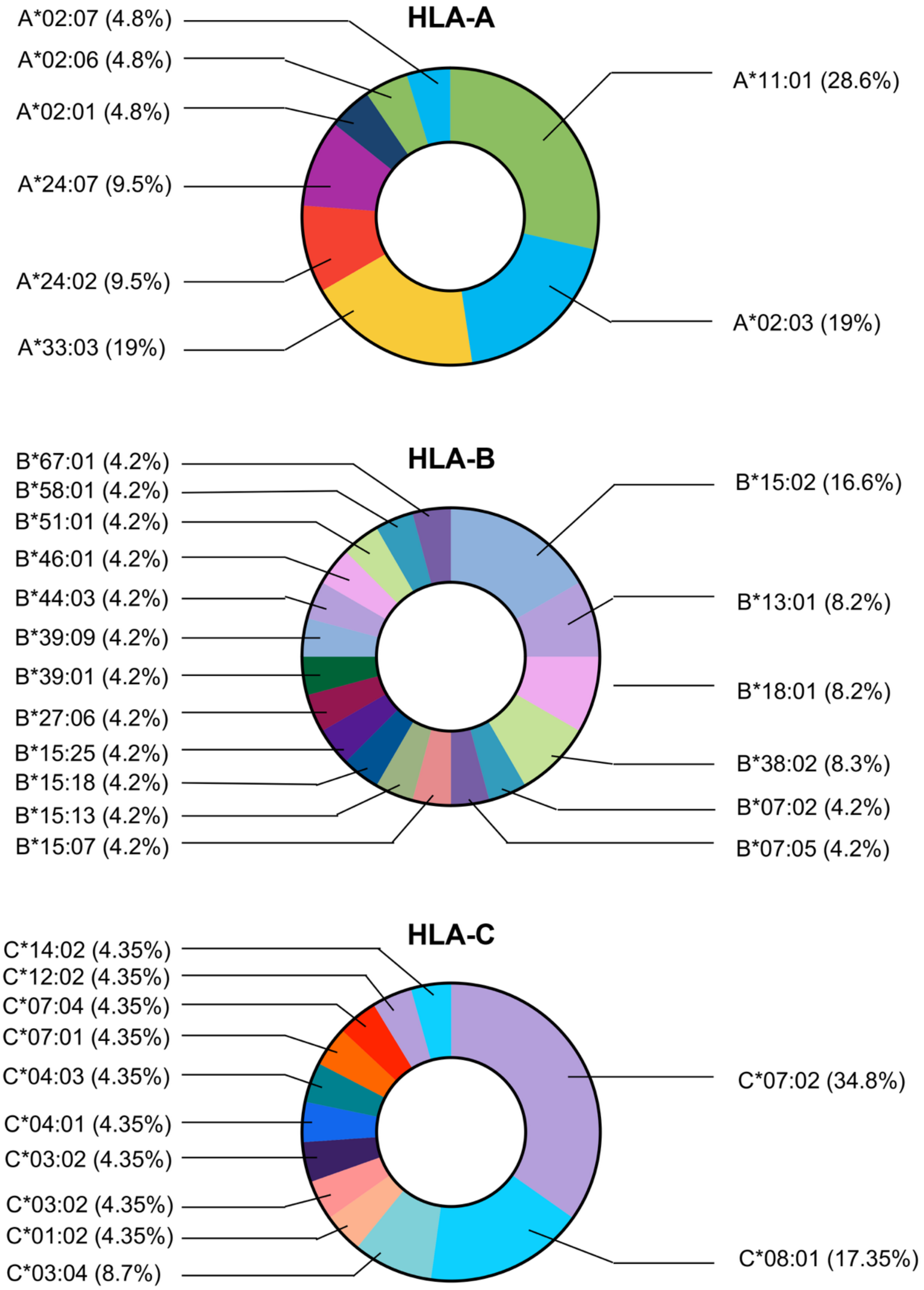


**Supplementary Figure S2: HLA distribution**

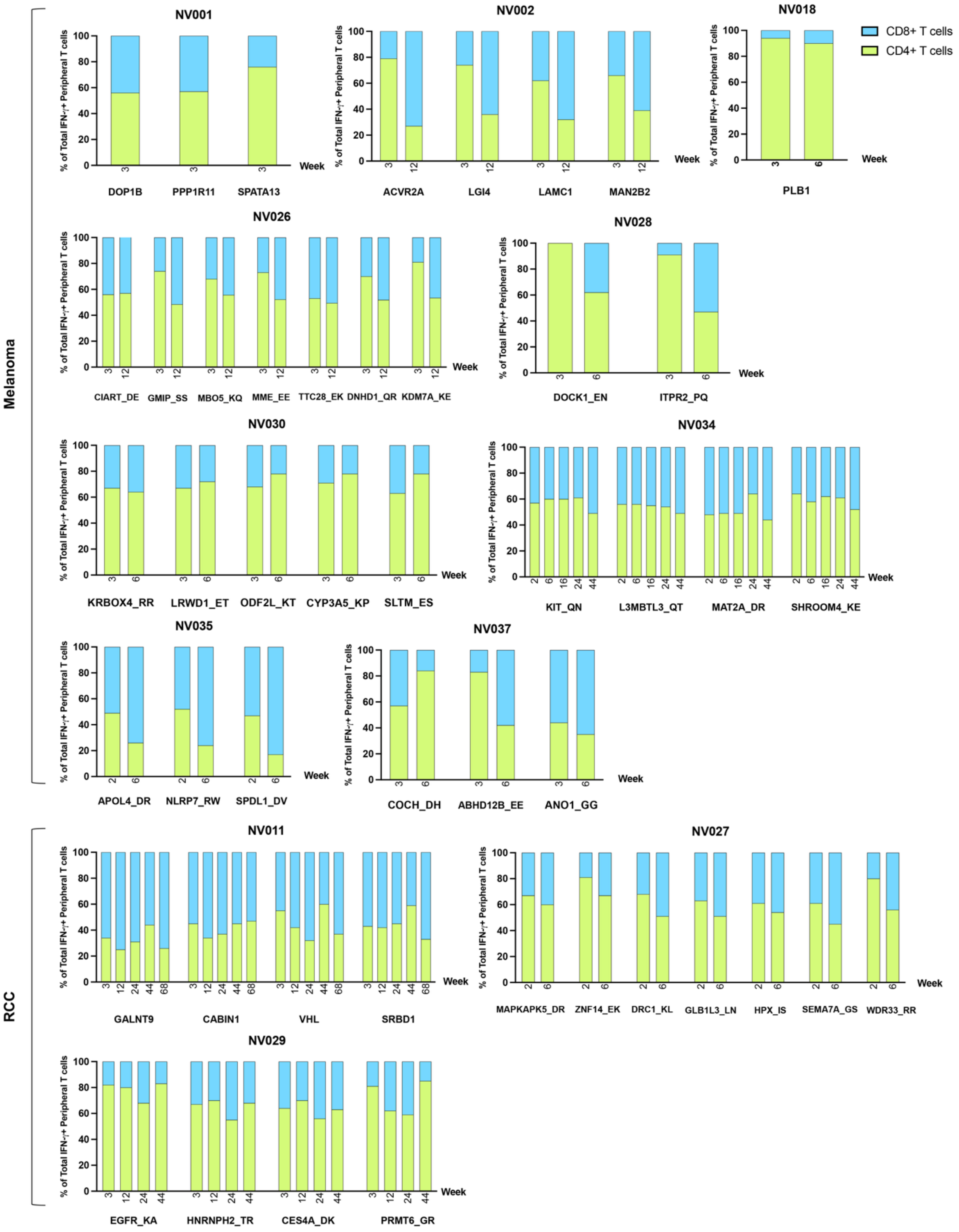


**Supplementary Figure S3 Characterization of neoantigen-specific CD4⁺ and CD8⁺ T-cell responses (all patients – CD4+ and CD8+ cells)**

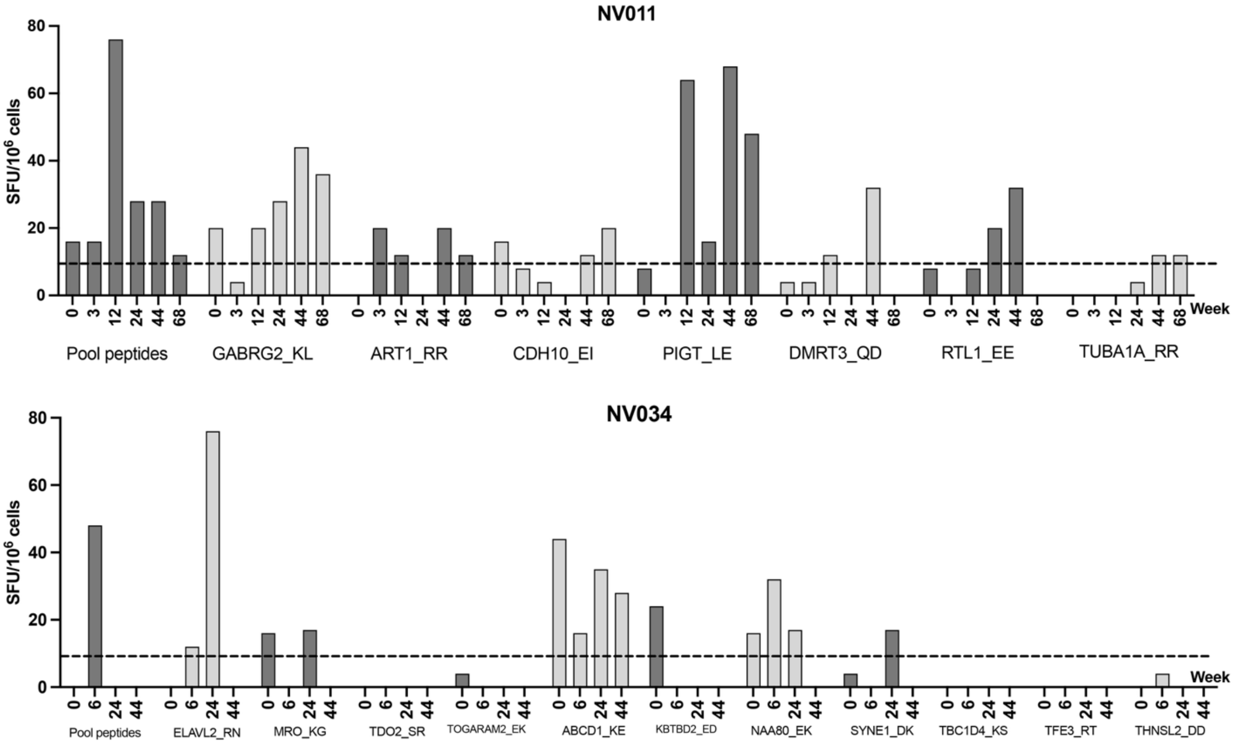


**Supplementary Figure S4: Evidence of epitope spreading in patient NV011.**
IFN-γ ELISpot responses in patient NV011 showing the emergence of T-cell reactivity to neoantigen peptides that were not originally included in the vaccine formulation. These de novo responses, absent at baseline, appear at later time points during the booster phase, indicating a broadening of the anti-tumor immune response beyond the initial vaccine targets.

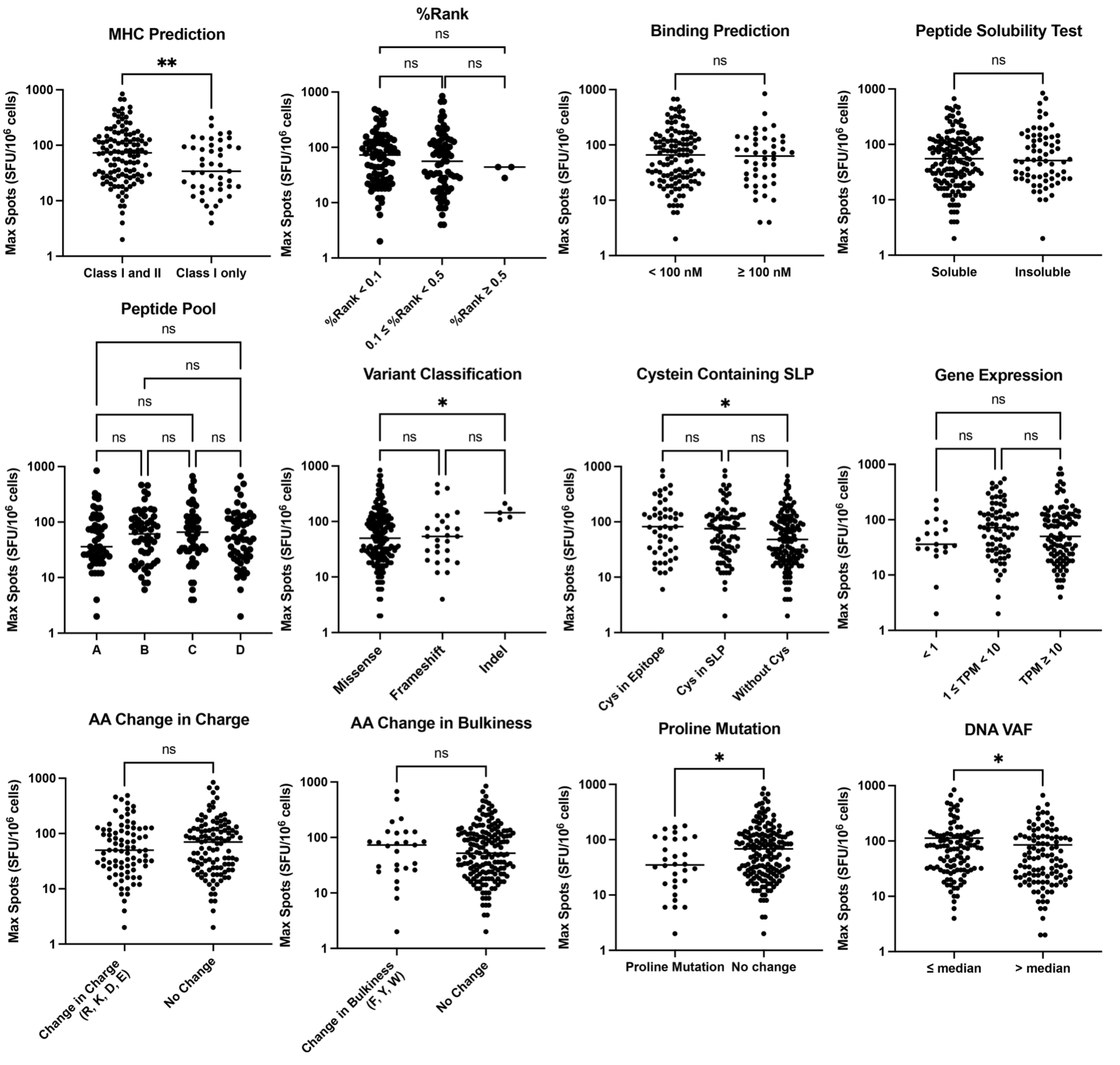


**Supplementary Figure S5: Analysis of factors contributing to peptide immunogenicity.** Plots showing the correlation between various peptide characteristics and immunogenicity, as measured by maximum ELISpot response. The analysis includes: binding prediction and % rank to both MHC Class I and II versus Class I alone; mutations involving proline and cysteine; and classification of the somatic variant (missense, frameshift, or indel). The data show that peptides with both MHC Class I and II predictions and those derived from indels were significantly more immunogenic, while peptides with proline mutations were less immunogenic. For statistical analysis of two groups, Mann-Whitney U test was used, while for three or more groups, Kruskal Wallis test with Dunn's multiple correction was applied (ns = non-significant, * $p<0.05$, ** $p<0.01$).
